# A specific, stable, and accessible LAMP assay targeting the HSP70 gene of *Trypanosoma cruzi*

**DOI:** 10.1101/2025.01.12.25320185

**Authors:** Sneider Alexander Gutierrez Guarnizo, Luciana Basma, Shirley Equilia, Beth Jessy Condori, Edith Malaga, Siena Defazio, Emily Arteaga, Jean Karla Velarde, Martín Obregón, Anshule Takyar, Carolina Duque, Jill Hakim, Freddy Tinajeros, Robert H Gilman, Natalie Bowman, Monica R. Mugnier

**Affiliations:** Department of International Health, Johns Hopkins University Bloomberg School of Public Health, Baltimore, MD, USA; Hospital Percy Boland Rodríguez, Ministerio de Salud Bolivia, Santa Cruz, Bolivia; Infectious Diseases Research Laboratory, Department of Cellular and Molecular Sciences, Universidad Peruana Cayetano Heredia, Lima, Perú; Molecular Microbiology and Immunology, Johns Hopkins Bloomberg School of Public Health, Baltimore, Maryland, USA; Department of Pathology, Johns Hopkins School of Medicine, Baltimore, Maryland, USA; Division of Infectious Disease, School of Medicine, University of North Carolina at Chapel Hill, Chapel Hill, North Carolina

**Keywords:** Acute Chagas, molecular diagnosis, LAMP, cost-effective method, HSP70 gene, congenital transmission

## Abstract

Diagnostic delays prevent most Chagas disease patients from receiving timely therapy during the acute phase when treatment is effective. qPCR-based diagnostic methods provide high sensitivity during this phase but require specialized equipment and complex protocols. More simple and cost-effective tools are urgently needed to optimize early Chagas disease diagnosis in low-income endemic regions. Here, we present a loop-mediated isothermal amplification (LAMP) that targets a highly conserved region in the HSP70 gene of *Trypanosoma cruzi,* the causative agent of Chagas disease. This assay demonstrates species-specific amplification across multiple parasite genetic lineages while maintaining stability after 2 hours of incubation and at least 8 months of storage at −20°C. Moreover, the assay is at least 12 times less expensive than the TaqMan qPCR that is currently routinely used for acute Chagas diagnostics. Population-based validation in 100 infants born to Chagas-positive mothers in Santa Cruz, Bolivia, yielded a specificity of 100% and sensitivity exceeding 77% when compared to a TaqMan qPCR that targets satellite DNA. This cost-effective assay holds promise for large-scale diagnosis of Chagas disease in endemic regions with limited resources.

## INTRODUCTION

*Trypanosoma cruzi*, the causative agent of Chagas disease, is endemic to the Americas, primarily affecting vulnerable populations and perpetuating poverty^1^. Annually, about 30,000 new cases are reported, with ∼6 million people estimated to be infected^2^. This devastating disease can cause severe clinical manifestations including megaesophagus, megacolon, and dilated cardiomyopathy, leading to an estimated 12,000 deaths per year^2^.

Chagas disease is transmitted through various routes, including vectorial (via triatomine bugs), oral, congenital, blood transfusion, organ transplantation, and lab accidents^3^. Despite successful vector control and blood screening, congenital transmission remains a key source of new *T. cruzi* infections, accounting for approximately 22% of cases and serving as the primary source of new infections in non-endemic regions^4,5^. Once infected, patients enter an acute phase characterized by high parasitemia^6^. During this phase, available treatments (benznidazole or nifurtimox) are highly effective and are well-tolerated in infants but can cause mild to severe side effects in adults^7,8^. The acute phase, especially in the context of congenital infection, offers a crucial window for intervention to prevent severe disease progression^7^. Unfortunately, the absence of symptoms or the presence of nonspecific ones often delays diagnosis. Consequently, it is estimated that fewer than 1% of people with Chagas disease receive treatment, largely due to delayed detection^9,10^.

Serological tests are routinely used to diagnose Chagas disease in the chronic phase, which is characterized by very low parasitemia^16^. These tests, however, which rely on the presence of anti-*T. cruzi* antibodies, cannot reliably detect acute infections, including congenital infections, where anti-parasite antibody levels are typically low. Therefore, molecular diagnosis represents an ideal strategy for acute phase infection when parasitemia is high and infection is treatable^11–15^. The most common molecular diagnostics for acute phase infection are qPCRs targeting nuclear satellite DNA (SatDNA)^12,17,18^, but these are expensive, show variable sensitivity, and require sophisticated equipment that is often unavailable in the low- and middle-income regions affected by Chagas disease ^19,20^. This highlights the urgent need for cost-effective, robust, and simple DNA-based diagnostic assays. Without such tools, timely diagnosis of congenital Chagas and other acute forms of the disease will remain challenging, leaving infections untreated and increasing the risk of irreversible chronic Chagas complications^21,22^.

To help address the limitations of existing molecular diagnostics, two major isothermal DNA amplification technologies have emerged as alternatives to standard qPCR: loop-mediated isothermal amplification (LAMP), introduced in 2000^23^, and recombinase polymerase amplification (RPA), introduced in 2006^24^. Though it utilizes a more complex primer design, LAMP is notably less expensive than RPA^25^ and has been widely used in diagnosing other infectious diseases such as COVID-19 and malaria, demonstrating its potential as a tool for population level diagnosis^26–28^. Additionally, compared to qPCR, LAMP requires less hardware and is more tolerant of amplification inhibitors^29^.

Although LAMP assays have been developed for Chagas diagnosis previously^30–33^, existing assays have shown some limitations, including false positives^30^ and the inability to discriminate between infection with *T. cruzi* and infection with other trypanosomatids^31,34^. Additionally, there is some evidence that the extreme genetic variability of *T. cruzi* strains might affect LAMP assay performance^35^. The limitations of existing LAMP assays highlight the need for alternative targets.

A promising alternative is to target the conserved protein-coding genes of *T. cruzi*, which have yet to be explored in LAMP assays for Chagas disease. Protein-coding genes are particularly appealing because of their greater genetic stability, a critical factor for reliable primer alignment^36^. While often overlooked in diagnostics due to their low copy numbers, protein-coding genes in *T. cruzi* often exist as multigene families, affording both the high copy numbers required for test sensitivity as well as the sequence conservation needed to develop a sufficiently specific diagnostic.

In this study, we developed a novel LAMP assay targeting a highly conserved region in the multicopy, protein-coding *HSP70* gene family of *T. cruzi*. Our LAMP protocol successfully amplified *T. cruzi* DNA from five distinct parasite genetic lineages without amplifying DNA from the related parasites *T. rangeli* and *Leishmania spp.* The assay proved robust and stable under various conditions, including a variety of incubation times and long-term storage at −20°C. It also produced consistent results across various sample types. Evaluating congenital Chagas in a population of Santa Cruz Bolivia, the assay showed optimal specificity and high sensitivity. This portable method could enable large-scale diagnosis and patients monitoring in endemic regions lacking economic or infrastructural resources.

## RESULTS

The *T. cruzi HSP70* subfamily genes, which serve as molecular chaperones involved in protein folding and transport, are an appealing target for a LAMP assay. In *T. cruzi,* there are an average of 88 *HSP70* genes per genome (TriTryp DB, Release 68). Moreover, the gene appears to be highly species-specific, in terms of both organization and sequence, potentially allowing for the specific discrimination of *T. cruzi* from other trypanosomatid parasites^37,38^. To assess the potential of *T. cruzi HSP70* as a diagnostic target, we performed a Neighbor-Joining tree analysis to compare the *HSP70* sequences across various *T. cruzi* strains (n=36) and two closely related trypanosomatids, *Leishmania* spp. (n=11), and *T. rangeli* (n=4) which infect humans in the same geographic region as *T. cruzi*. The *T. cruzi HSP70* sequences cluster in a distinct clade compared to the *HSP70* of *T. rangeli* and *Leishmania* spp. (**Fig. 1, Supplementary File 1**). The *T. cruzi HSP70* sequences exhibited a high identity of 98.63% (**Supplementary File 1)**, indicating that this sequence is highly conserved and serves as a feasible target for designing LAMP primers.

**Figure 1.**
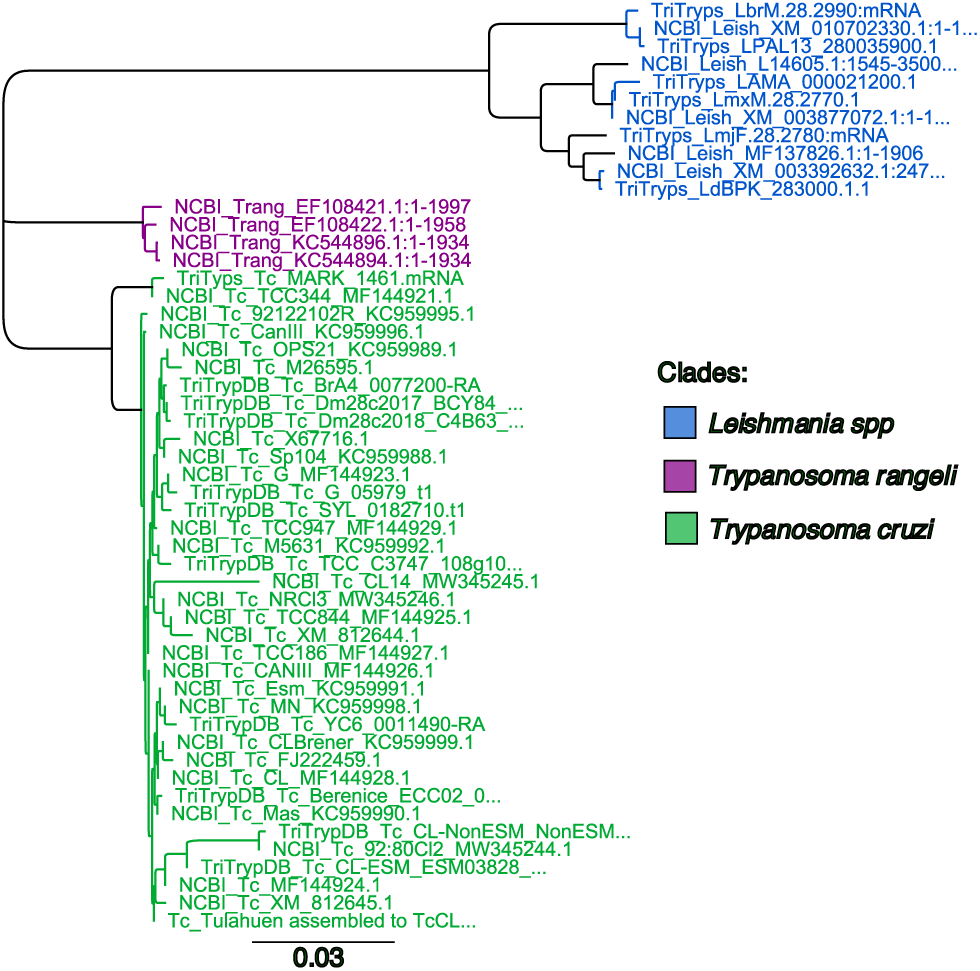
The *HSP70* sequence is a molecular marker that differentiates *T. cruzi* from closely related trypanosomatids. Neighbor-joining tree analysis based on *HSP70* sequences discriminates *Leishmania* spp (blue), *T. rageli* (purple), and *T. cruzi* (green). The horizontal scale represents the evolutionary distance. A scale of 0.03 means an average of 3 substitutions per 100 nucleotides.

Next, the LAMP*_TcHSP70_* primers were designed along a region of 236 bp at the end of the *HSP70* open reading frame. This region was conserved across *T. cruzi* strains but included sequence variations in *Leishmania* spp. and *T. rangeli* to minimize primer binding to these other species. Eight subregions were defined (F3, F2, FLc, F1, B1c, BL, B2c, B3c), resulting in six primers, the forward outer primer (F3), the backward outer primer (B3), the forward loop primer (FL), the backward loop primer (BL), the forward inner primer (FIP), and the backward inner primer (BIP). The FIP and BIP primers create the forward and backward loop, respectively, and align in two regions each. FIP aligns in F2 and F1c while BIP aligns in B2c and B1c (**Figure 2, (Supplementary File 2-3)**. The designed primers showed Gibbs free energy (ΔG) values between −3.61 and −11.49, with primers F3 and F1c showing moderate risk for self and heterodimers **(Supplementary File 4).**

**Figure 2.**
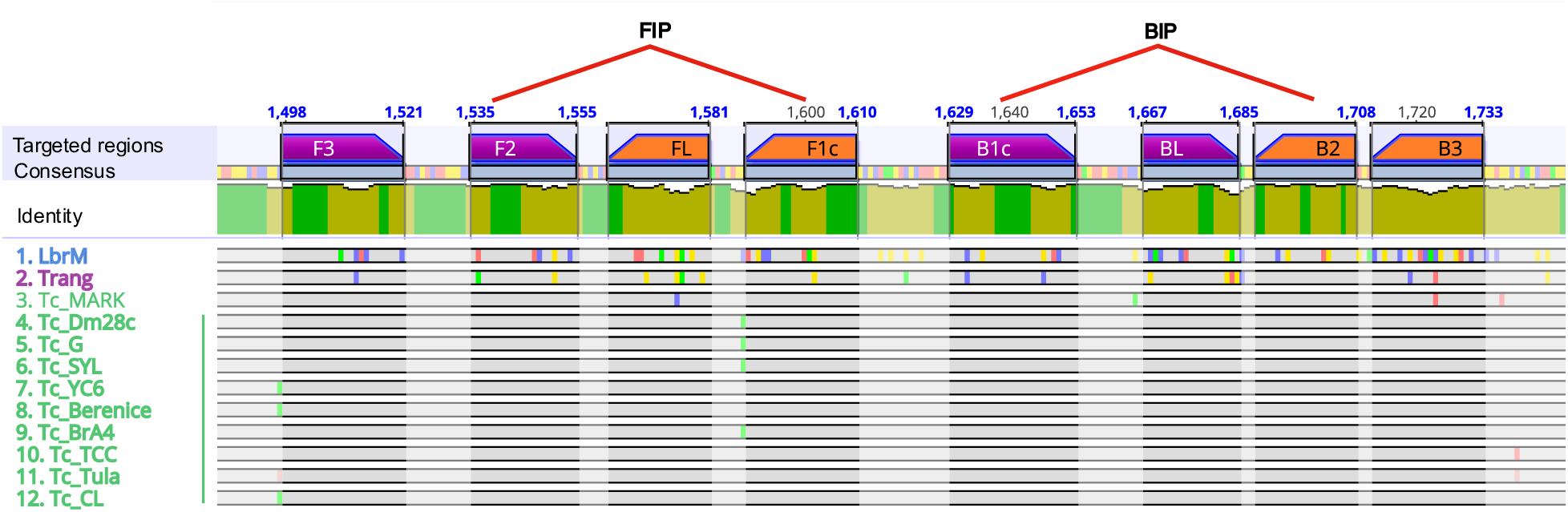
Primers for LAMP*_TcHSP70_* align to regions that distinguish *T. cruzi* from *Leishmania* and *T. rangeli*. The HSP70 sequences from twelve representative strains of *Leishmania*, *T. rangeli*, and *T. cruzi* were aligned using the Clustal Omega algorithm. The regions where the primers align in forward and reverse are shown as purple and orange bars, respectively. The inner primers (FIP and BIP) align to two distinct regions, connected by red lines. Nucleotide variants are depicted as small colored bars over the sequences represented as horizontal grey bars. All *T. cruzi* sequences, except *T. cruzi* marinkellei, which does not infect humans, show 100% identity in the primer-targeted regions. The figure was generated using Geneious Prime software. Sequences named on the left: 1) *Leishmania braziliensis*, 2) *T. rangeli*, 3) *T. cruzi* marinkellei, 4) *T. cruzi* Dm28c, 5) *T. cruzi* G, 6) *T. cruzi* Sylvio, 7) *T. cruzi* YC6, 8) *T. cruzi* Berenice, 9) *T. cruzi* Brasil A4, 10) *T. cruzi* TCC, 11) *T. cruzi* Tulahuen, 12) *T. cruzi* CL.

To test the LAMP*_TcHSP70_* primers, we conducted a preliminary reaction evaluating the amplification under the conditions established by Ordóñez et al., 2020^33^. This involved incubating for 60 minutes at 65°C and adapting it to the SYBR Green-based One-pot system^39^ (**Figure 3A**). We evaluated two representative *T. cruzi* strains (Sylvio and G) and two negative controls (no template control and human DNA control). A positive result was visible as a color change from orange to green under visible light or as green fluorescence under UV light (**Figure 3B**). The results were confirmed by running on an agarose gel which showed amplification bands for both *T. cruzi* strains and no bands for the negative controls. Additionally, the agarose gel did not show significant formation of primer dimers (**Figure 3C**).

**Figure 3.**
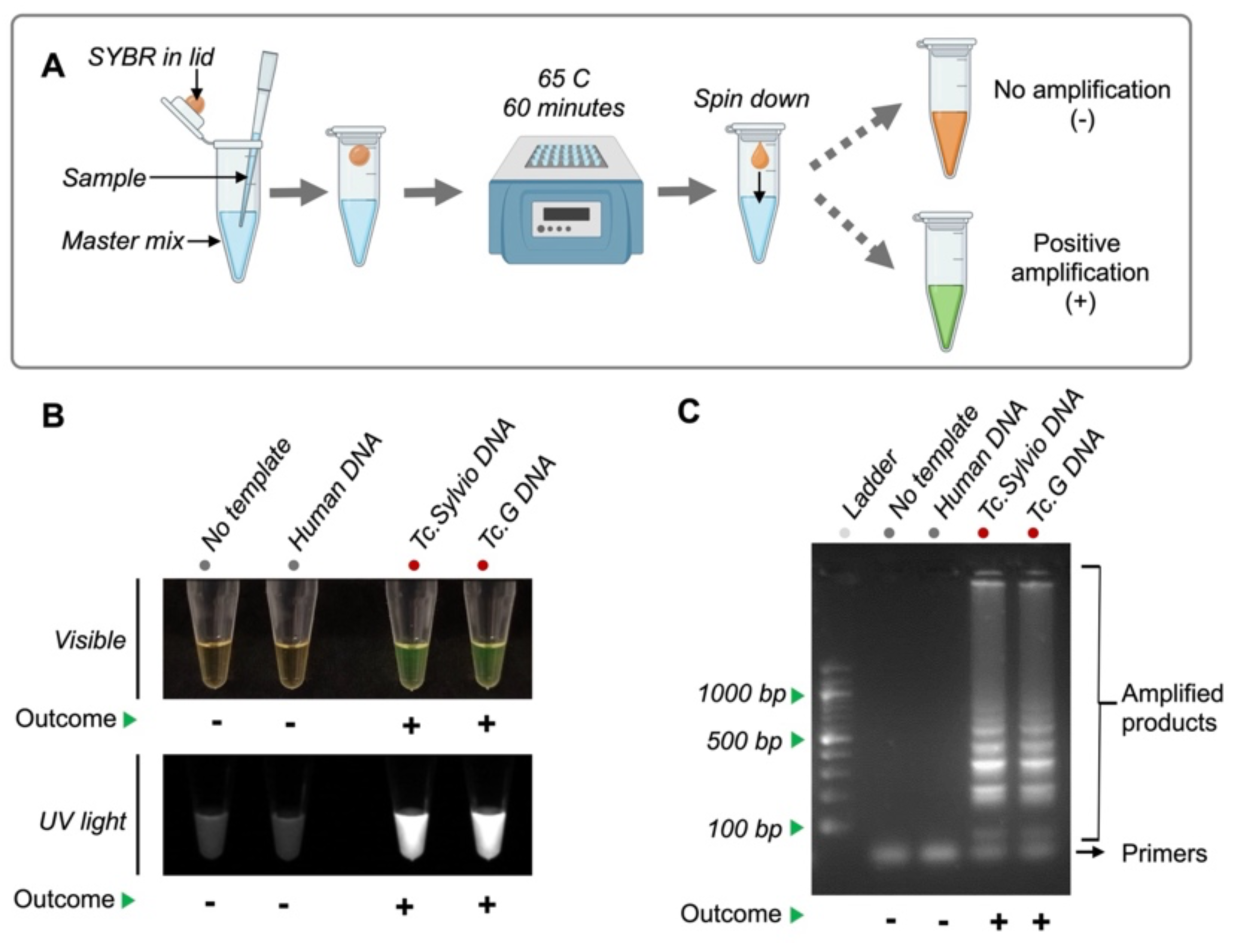
LAMP*_TcHSP70_* amplifies *T. cruzi* DNA. **A)** Workflow for LAMP*_TcHSP70_* amplification. **B)** Visualization of the amplified products by visible light and UV light. 1ng DNA was loaded per sample. **C)** Visualization of the amplified products by electrophoresis. Representative agarose gel of three replicates showing the classic pattern of LAMP product. 3 μL of the amplified products were visualized on a 2% agarose gel pre-stained with 0.2 μg/mL ethidium bromide. A 100 bp DNA ladder was used as a reference for determining amplicon sizes.

Next, we evaluated the specificity of the LAMP*_TcHSP70_* assay for *T. cruzi* and its ability to amplify various distinct strains. We tested 40 samples, including 2 negative controls, 2 *Leishmania* species, *T. brucei, T, rangeli,* and 34 *T. cruzi* strains. The LAMP*_TcHSP70_* did not amplify DNA from any species other than *T. cruzi*. The LAMP*_TcHSP70_* showed a preliminary sensitivity of 97% (95% CI: 85% to 100%), and a specificity of 100% (95% CI:40% to 100%). Only the isolate *T. cruzi* Mg TcI from Colombia did not show positive amplification, possibly indicating genetic variability in the *HSP70* gene in this strain (**Figure 4**).

**Figure 4.**
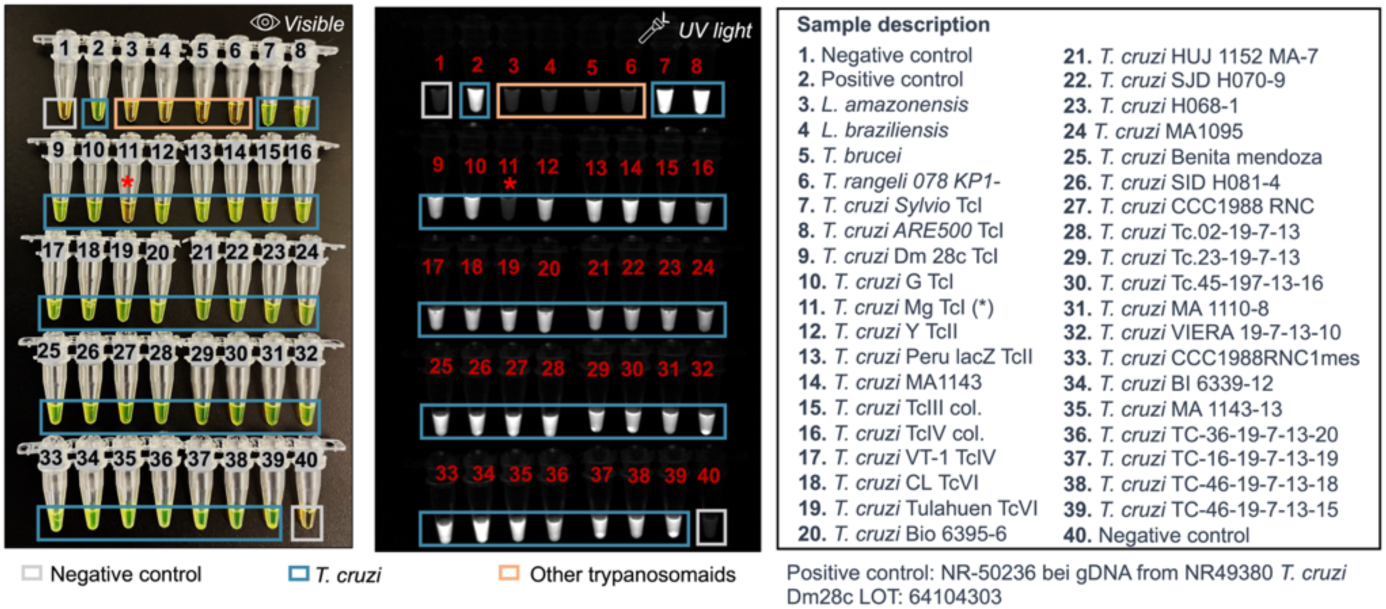
LAMP*_TcHSP70_* shows high specificity and sensitivity using parasite-derived DNA. 40 samples were run in parallel. Representative captures of two replicates evaluated by visible and UV light. For all samples, 1 ng of purified DNA was seeded.

After confirming that the LAMP*_TcHSP70_* has good specificity and promising sensitivity, we assessed the analytical limit of detection (LoD). To achieve this, we serially diluted *T. cruzi* DNA to identify the lowest DNA amount yielding a positive amplification. We evaluated seven reference strains representing the five different *T.cruzi* lineages known as discrete typing units (DTUs). DTUs broadly represent the parasite’s genomic variability. Initial tenfold serial dilutions established a preliminary LoD of 0.01 ng (**Fig. 5A-B**). To refine this estimate, we conducted further twofold dilutions for two representative strains, resulting in a final LoD of 1.25 pg. The LoD was consistent across different *T. cruzi* lineages and is equivalent to 0.068 ± 0.032 parasites (**Fig. 5C-D, Supplementary File 5-6**). When comparing the LAMP*_TcHSP70_* to the routine TaqMan SatDNA qPCR used for acute Chagas diagnosis^40^, the LAMP*_TcHSP70_* was able to detect DNA in samples with a SatDNA qPCR Ct value up to 28.37± 0.26 (**Supplementary File 7**). These results suggest that LAMP*_TcHSP70_* might have optimal sensitivity for detecting acute Chagas infections characterized by high parasitemia.

**Figure 5.**
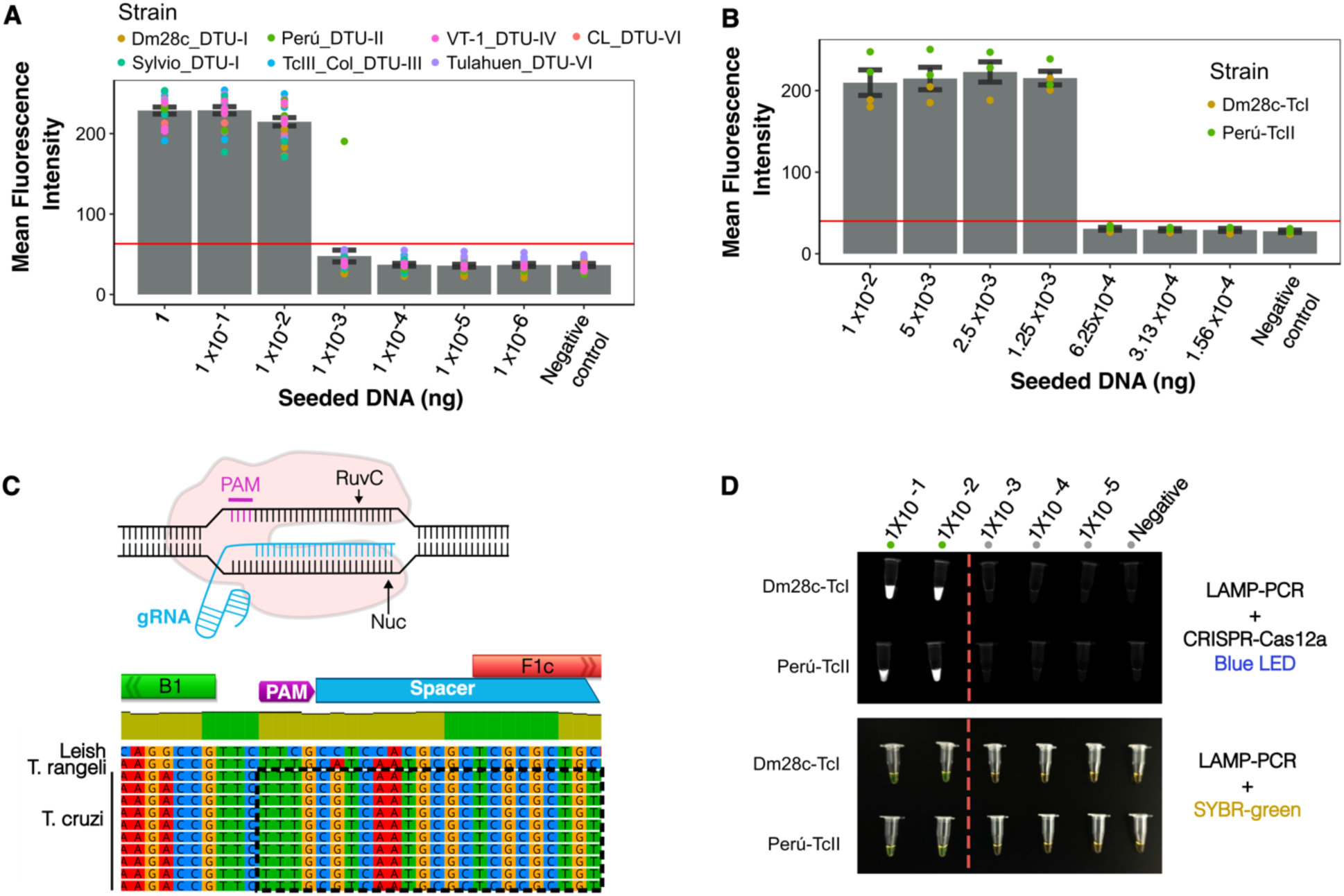
The LAMP*_TcHSP70_* assay shows a consistent LoD across multiple strains and DTUs, with a LoD in the order of picograms. **A)** <u>LoD</u> after the first round of serial DNA dilutions using a factor of 10. **B)** <u>LoD</u> after the first round of serial DNA dilutions using a factor of 2. **C)** Design of CRIPRcas12a system. **D)** <u>LoD ofter using</u> LAMP*_TcHSP70_* coupled to CRIPRcas12a system. Three independent replicates per strain. Mean fluorescence intensity was estimated using the ROI manager tool of Fiji 2 software. A-B) Error bars indicate standard error. The horizontal red line highlights the mean fluorescence intensity reached by the negative control.

We next evaluated whether the addition of CRISPR-Cas12a to the LAMP*_TcHSP70_* could improve the LoD as demonstrated in previous studies targeting different pathogens^39,41–44^. We designed a gRNA to specifically recognize a sequence between the B1 and F1c regions, where the inner primers align to form the loops (**Figure 5C**). As shown in **Figure 5D**, although the CRISPR-Cas12a system improved the discrimination between positive and negative amplification, it did not significantly extend the LoD. Our cost analysis revealed that the standard TaqMan SatDNA PCR is 14.3 times more expensive than LAMP with SYBR Green and 12.9 times more expensive than LAMP with the Cas12a system, highlighting the cost-effectiveness of the LAMP approach (**Supplementary File 8**).

Once the LoD for our system was established, we then assessed the minimum incubation time required for optimal amplification of 2pg of DNA of the template. Our findings demonstrate that incubation at 65°C for 60 minutes is needed to detect low abundance targets (**Fig. 6A**). Although previous studies have suggested that LAMP reaction time should not exceed 30 minutes to prevent inaccurate results^45,46^, our LAMP targeting *T.cruzi HSP70* proved highly specific and stable, as no nonspecific amplification was observed after 120 minutes of incubation (**Fig. 6A, Supplementary File 9**). We then evaluated the stability of the complete master mix at −20°C over time. After 8 months, the master mix showed no significant difference in mean fluorescence intensity (**Fig. 6B, Supplementary File 9**). These results indicate that LAMP*_TcHSP70_* is stable for prolonged incubation and storage. The reaction can thus be prepared in large batches to streamline daily sample processing.

**Figure 6.**
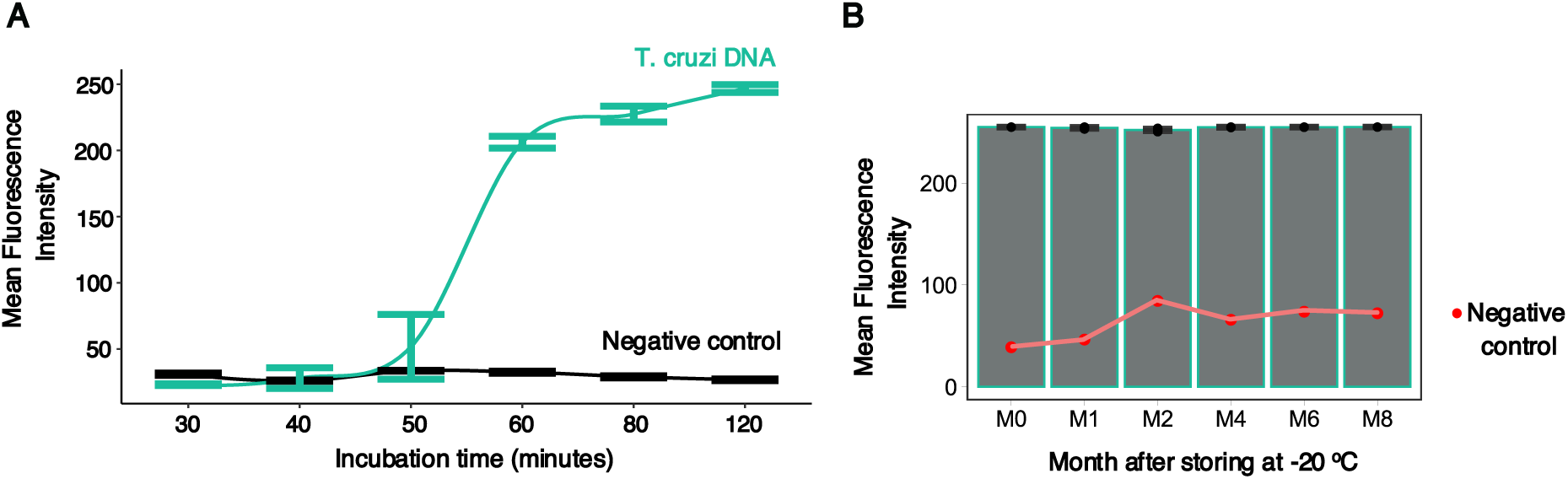
The LAMP targeting *T.cruzi HSP70* remains stable even after extended incubation and storage times. **A)** Extended incubation times do not reveal non-specific amplification in negative controls. Data represent four technical replicates per incubation period. Human DNA (negative control) was seeded at 50 ng, while *T. cruzi* DNA was seeded at 2 pg. **B)** Master mix storing for eight months does not affect LAMP performance. Data represent three technical replicates per storing period. One reaction was used for negative control. Human DNA (negative control) was seeded at 50 ng, while *T. cruzi* DNA was seeded at 1 ng. Error bars indicate standard error. Mean fluorescence intensity was estimated by ROI area using Fiji software.

Having demonstrated good specificity, sensitivity, and stability of our LAMP*_TcHSP70_* assay with parasite isolates, we then evaluated its performance with clinical samples. A total of 100 infants born to Chagas-positive mothers were analyzed, for each infant, DNA was extracted from peripheral and/or umbilical cord blood. Ninety-two infants had a single sample tested, while eight infants (Infant IDs: 1, 2, 15, 19, 27, 31, 34, 76) had more than one sample evaluated resulting in a total of 109 samples (**Supplementary File 10)**. We compared both the sample-based and patient-based performance of LAMP*_TcHSP70_* with the TaqMan-based SatDNA-qPCR, the routine diagnostic test for congenital Chagas disease, using a blinded approach. For the 109 samples evaluated, LAMP demonstrated a sensitivity of 77.27% (95% CI: 54.63% to 92.18%) identifying 17 out of 22 positive samples by qPCR, a specificity of 100% (95% CI: 95.85% to 100.00%), and an accuracy of 98.86% (94.63% to 99.95%) (**Fig 7A**). When analyzing results per patient, where at least one positive sample classified the infant as positive, LAMP showed a sensitivity of 87.5% (95% CI: 61.65% to 98.45%) identifying 14 out of 16 positives by qPCR, a specificity of 100% (95% CI: 95.7% to 100.00%), and an accuracy of 99.37% (95% CI: 95.2% to 100%) (**Fig 7B)**. Notably, LAMP*_TcHSP70_* performed better with samples with high parasitemia but also detected samples with low parasitemia as shown in **Figure 7C**. These findings suggest that LAMP*_TcHSP70_* is a viable alternative to SatDNA-qPCR for diagnosing acute Chagas, especially in resource-limited settings.

**Figure 7.**
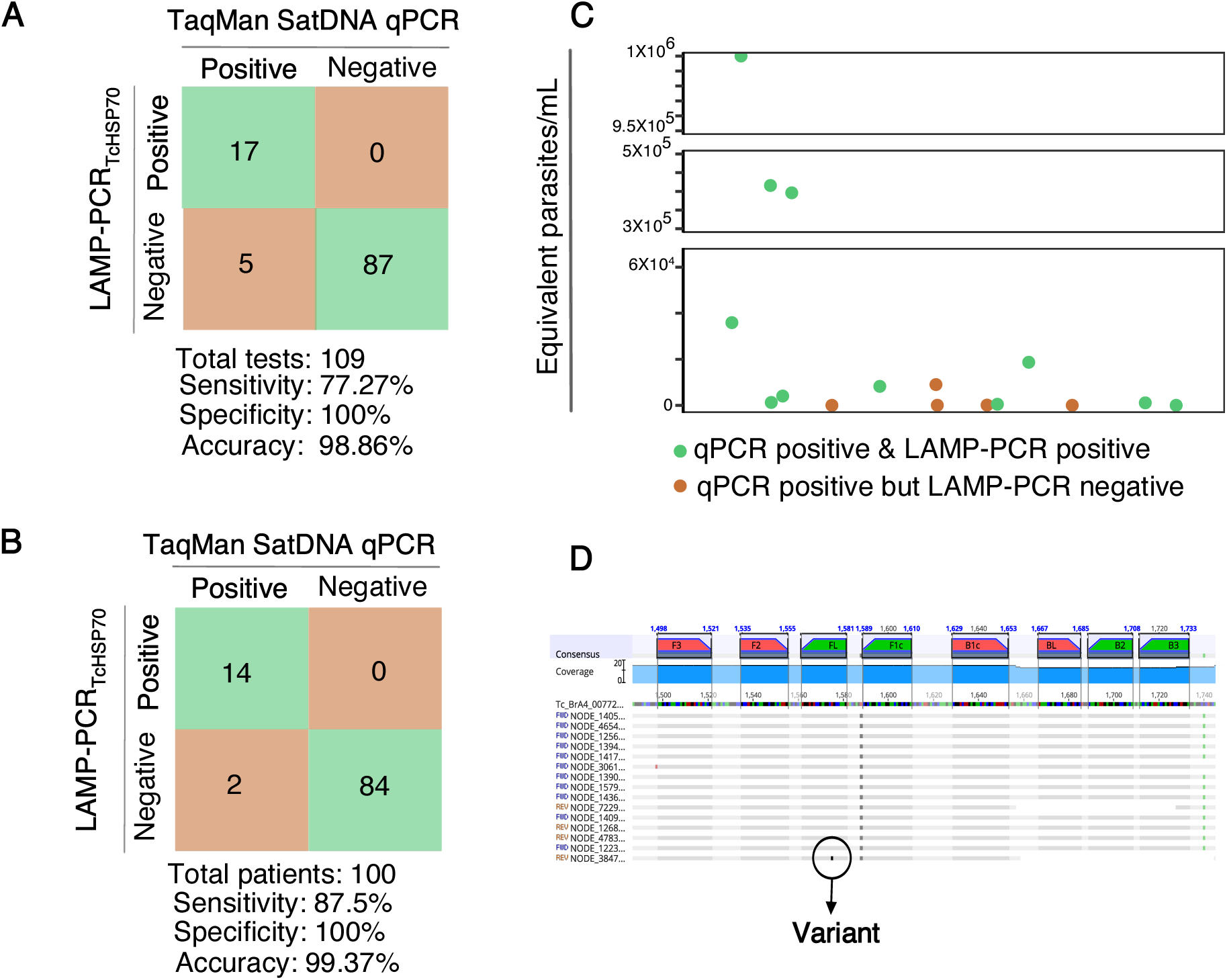
The LAMP*_TcHsp70_* assay exhibits high specificity and accuracy but lower sensitivity compared to SatDNA-qPCR. **A)** Sensitivity, specificity, and accuracy per test. **B)** Sensitivity, specificity, and accuracy per patient diagnosis. **C)** Positive and negative LAMP*_TcHSP70_* results in relation to parasitic load (parasites/mL) as estimated by the qPCR calibration curve. **D)** Alignment of *HSP70* sequences from clinical isolates.

Besides sensitivity, the failure of LAMP to amplify certain samples may also stem from issues with primer alignment. *HSP70* is a highly conserved gene, but *T. cruzi* parasites exhibit significant genomic plasticity. The inability of our LAMP to amplify certain samples detected by standard PCR (e.g., infant IDs: 4, 19) might also be the result of genomic variations, such as mutations or low gene dose, that affect LAMP performance. Additional DNA from the patients tested was unavailable to investigate *HSP70* sequence variations potentially impacting primer alignment, so we analyzed the *HSP70* sequence of 15 clinical *T. cruzi* isolates from additional Bolivian patients. The analysis confirmed that the *HSP70* sequence is generally highly conserved, revealing only one variant (p1575 T −> G). This variant does not affect the 3’ or 5’ primer ends, suggesting a low impact on primer performance (**Figure 7D**). However, these findings underscore that local parasite populations may harbor low-frequency variants that could impact primer alignment for the LAMP and other assays.

## DISCUSSION

Early diagnosis is critical for the treatment of Chagas disease but is hampered by the lack of a molecular gold standard and the high cost of traditional methods. Here we present a new specific, stable, and accessible LAMP assay based on the detection of *T. cruzi HSP70* gene.

LAMP has emerged as a cost-effective method for expanding the use of molecular diagnostics at the population level^25^, yet only a few assays exist for detecting *T. cruzi*. Primarily focused on established qPCR targets, these assays show variable sensitivity (69.2%-97%)^30–32^ and frequently suffer from nonspecific amplification resulting in false positives^30^ or an inability to distinguish *T. cruzi* infection from infection with related trypanosomatids^31,34^ leading to a reduced specificity. Moreover, the primer sequences for the most promising SatDNA-targeting LAMP have not been made publicly available, limiting its use and independent validation^30,32,47^. Given these limitations, we sought to develop an open-access LAMP assay targeting a novel highly specific *T. cruzi* sequence.

Our phylogenetic analysis confirmed the effectiveness of *HSP70* in distinguishing *T. cruzi* from related species, enabling the design of LAMP primers targeting eight highly conserved regions. Using this primer set, we found that our LAMP*_TcHSP70_* assay demonstrated 100% specificity when tested with reference strains and clinical samples, with no false positives even after doubling the incubation time. This indicates a lack of non-specific primer interactions, a common issue in LAMP assays. Furthermore, unlike 18S rRNA targeting assays, our HSP70-based assay avoids cross-amplification with *Leishmania*, further highlighting its high specificity. Additionally, we showed that the LAMP*_TcHSP70_* master mix is extremely stable long term, facilitating large batch preparations and streamlining master mix preparation.

Despite having a lower copy number compared to SatDNA, kDNA, or 18S rRNA, the *HSP70* targeted by our LAMP demonstrates high analytical sensitivity with a limit of detection equivalent to less than 0.07 parasites per reaction. This sensitivity is reasonable for diagnosing acute Chagas, especially congenital Chagas which is characterized by high parasitemia particularly after the firth month of birth. After analyzing clinical samples, the LAMP*_TcHsp70_* showed a sensitivity of 87.5%. This somewhat reduced sensitivity relative to existing qPCR assays is likely the result of differences in target copy number: while parasites harbor an average of 88 copies of *HSP70* per genome, they harbor 10^4^ to 10^5^ copies of the DNA satellite region^48^. Although we cannot rule out the fact that mutations affected the assay performance, our evidence suggests *HSP70* mutations are unlikely to be a major problem. Sequencing of clinical isolates from Santa Cruz, Bolivia, did not reveal any mutations that would interfere with primer amplification. *T. cruzi* reproduces asexually, primarily through binary fission, so we would not anticipate rapid accumulation of mutations in this gene under normal conditions. Nevertheless, our understanding of the genetic diversity of *T. cruzi* in the clinical setting is limited, and further sequencing of genomes from clinical samples will be needed to ensure the optimal design of LAMP primers and to clarify whether different strains have copy number variations in the HSP70 gene that might affect the sensitivity of the LAMPTcHsp70 assay. Another explanation for the lower sensitivity of LAMP*_TcHsp70_* when compared to qPCR is the possibility that some samples contained LAMP inhibitors. Notably, in this study, we used the Bst 2.0 polymerase, a reliable enzyme for DNA amplification in LAMP assays. The advent of more advanced polymerases, such as Bst 3.0, holds promise for improved sensitivity in future versions of the LAMP*_TcHSP70_* assay. This newer enzyme, which amplifies not only DNA but also RNA templates, could significantly enhance both the sensitivity and reaction speed of the assay.

Because our assay failed to detect *T. cruzi* DNA in 2/16 infants, we explored the integration of a CRISPR-Cas12a system to improve the assay’s sensitivity. Though it did not improve sensitivity, CRISPR-Cas12a did improve the signal-to-noise ratio of the assay, with a more refined readout compared to SYBR Green, which interacts non-specifically with any DNA in the reaction. Thus, the incorporation of the Cas12a system could be valuable in circumstances where samples are suffering from high background amplification.

Regardless of the detection method (SYBR Green or Cas12a), our LAMP*_TcHSP70_* remains at least 12 times cheaper than traditional qPCR assays. Moreover, we have made the entire LAMP*_TcHSP70_* protocol publicly available. The assay’s ease of use, low cost, and open protocol not only facilitate independent validation of the method but also provide an option for large-scale, low-cost screening of suspected acute Chagas infection.

Such large-scale screening is particularly critical in the context of congenital Chagas, an especially important avenue for control of Chagas disease. Approximately 22% of new cases are congenitally transmitted, and congenital infection—an acute infection—is uniquely treatable^5^. When diagnosed and treated within the first year of life, congenital *T. cruzi* infection has a >90% cure rate^4^. Therefore, large-scale diagnosis of newborns in endemic regions could significantly increase the number of detected, and thus treated, cases of acute Chagas. Such population-level diagnosis has been infeasible until now, for reasons related to both logistics and cost. LAMP*_TcHSP70_* overcomes these challenges, being stable, specific, and exhibiting good sensitivity when applied to clinical samples. Further efforts will be needed to adapt simplified and cost-effective DNA extraction protocols that facilitate sample processing in low-resource settings.

This study opens new avenues for analyzing novel targets to detect acute Chagas disease. LAMP continues to be a promising, cost-effective alternative to expand Chagas disease diagnosis and treatment at the population level in low-income regions.

## MATERIALS AND METHODS

### DNA and strains of reference

We used DNA from reference strains obtained from ATCC and BEI resources, including *T. cruzi* strains representative of DTU I (Dm28c NR-49380, Sylvio-X10 ATCC-50823, G NR-49382), DTU II (Y NR-46429, Peru LacZ Clone 4 NR-18960), DTU IV (TcVT-1 NR-46428), and DTU VI (Tulahuen LacZ clone C4 NR-18959, CL). Additionally, we used DNA from *L. amazonensis* (NR-49247) and *L. braziliensis* (NR-50608). DNA from *T. cruzi* Mg Tc I, *T. cruzi* TcIII col, and *T. cruzi* TcIV col strains were provided by Dr. Juan David Ramirez from Universidad del Rosario. DNA from the reference strain *T. rangeli* 078 KP1- was donated by Dr. Daniel Urrea from the LIPT laboratory at Universidad del Tolima. All remaining isolates were obtained from Dr. Gilman’s repository (IRB00007176).

### Population of study

Sample collection was approved by the Institutional Review Boards of the University of North Carolina at Chapel Hill (IRB 19–3014) and Hospital De La Mujer Dr. Percy Boland (Protocol 036). Pregnant women were recruited at Hospital De La Mujer Dr. Percy Boland in Santa Cruz, Bolivia, over 12 months between June of 2023 and June of 2024. A total of 1,344 mothers were enrolled, 252 of whom were seropositive for Chagas disease. Umbilical cord blood samples were collected at birth, and infant peripheral blood was obtained post-delivery. Based on routine TaqMan SatDNA qPCR, 16 infants were diagnosed with congenital Chagas. This study evaluated samples from 100 infants, including 84 controls and 16 cases of congenital Chagas that had sufficient DNA left over after performing the routine diagnostics. All samples/patient IDs were deidentified and not known to anyone outside the research group.

### Alignment of HSP-70 sequences

The *HSP70* sequence of *Trypanosoma cruzi* Brazil A4 (TcBrA4_0077200, TriTryps DB) was used as input for a BLAST search against *T. cruzi* strains, *Leishmania*, and *T. rangeli* strains available in the TriTryps and NCBI databases. Representative sequences with the highest identity were retained for each strain. A total of 51 representative sequences were evaluated: 36 from *T. cruzi*, 11 from *Leishmania*, and 4 from *T. rangeli*. The *HSP70* sequences were analyzed using a Neighbor-Joining tree with Geneious Prime® 2024.0.5 software. The parameters used were global alignment with free gaps, a cost matrix of 70% similarity (IUB) (5.0/-4.5), and the Tamura-Nei genetic distance model.

### DNA extraction

DNA from parasite pellets was purified using the QIAamp® DNA Blood Mini Kit (Catalog number or CN: 51104) according to the manufacturer’s instructions. DNA from patient samples was purified using the High Pure PCR Template Preparation kit (Roche Diagnostics GmbH, Mannheim, Germany; REF: 11796828001) as previously described^40^. An internal amplification control (IAC) was prepared at 40 pg/µL. For blood samples, 300 µL of blood was mixed with Guanidine/EDTA and 5 µL of IAC (40 ρg/µL). Additionally, 40 µL of Proteinase K (20 mg/mL) was added for lysis and incubated at 70°C for 10 minutes. All DNA samples were resuspended in 50 µL of DNAse/RNAse-free water.

### Validatory satDNA TaqMan qPCR

A duplex qPCR was conducted for all clinical samples following the protocol previously published by Mayta et al., 2019 ^40^. Briefly, the PCR targets the satellite sequence of the nuclear genome of *T*. *cruzi* and IAC. The qPCR was performed according to published methods, using primers Cruzi 1 (5’–ASTCGGCTGATCGTTTTCGA–3’) and Cruzi 2 (5’– AATTCCTCCAAGCAGCGGATA–3’) to amplify a 166-bp fragment of nuclear satellite DNA. The probe cruzi 3 (5’–CACACACTGGACACCAA–3’) was labeled with 5′FAM (6– carboxyfluorescein) and 3′MGB (minor groove binder)^40^.

### LAMP *_TcHSP70_* primer design

Upon sequence alignment, primers were manually designed in regions with high coverage and identity for *T. cruzi* strains. The primers were designed to maximize variants in *Leishmania* or *T. rangeli* homologus sequences. The likelihood of self-dimer and heterodimers were evaluated by the Oligoanalyzer IDT tool using default parameters.

### LAMP master mix and incubation

The master mix preparation was adapted from a protocol by Ordóñez et al., 2020^33^. The master mix was prepared in batches of 50-100 reactions in 23 µL and contained: 40 pmol FIP primer, 40 pmol BIP primer, 0.5 pmol F3 primer, 0.5 pmol B3 primer, 0.5 pmol LF primer, 0.5 pmol LB primer, 2.5 mM dNTPs (NEB, CN: N0447S), 0.8 mM MgSO₄ (NEB, CN: B1003S), 1 M Betaine (Sigma-Aldrich, CN: B0300-5VL), 1X Isothermal Amplification Buffer (20 mM Tris-HCl pH 8.8, NEB, CN: B0537S), and 8 U Bst Polymerase 2.0 WarmStart (NEB, CN: M0537L).

Two microliters of DNA template were added to the master mix to achieve a total volume of 25 µL. SYBR Green I dye (1:10 dilution) (Invitrogen, CN: S-7567) was then added to the tube lid of the tube. The reaction was incubated at 65°C for 60 minutes. After incubation, the vials were centrifuged to combine the dye with the amplified product.

### LAMP visualization

The amplified products were initially visualized by the eye using a black background. For quantitative analysis, the amplified products were imaged using an Azure Biosystems 200 gel imager with a 365 nm light source and a 595 nm emission filter. The mean fluorescence intensity was calculated using the ROI manager function of fiji2 software.

Additionally, during standardization, the amplified products were also visualized on agarose gel. The products were pre-incubated at 80°C for 10 minutes to deactivate the enzyme and stop the reaction. Then, 3 μL of the amplified products were visualized on a 2% agarose gel (Invitrogen, CN: 16-500-500), pre-stained with 0.2 μg/mL ethidium bromide. The electrophoresis was performed at 100V for 35 minutes.

### Analytical limit of detection LAMP*_TcHSP70_*

DNA from seven *T. cruzi* strains (Dm28c_DTU-I, Sylvio_DTU-I, Perú_DTU-II, TcIII_Col_DTU-III, VT-1_DTU-II, Tulahuen_DTU-VI, CL-DTU-VI) was purified and quantified using a Qubit fluorometer (ThermoFisher, CN: Q32850). The DNA concentrations were normalized to 0.5 ng/µL, resulting in a total of 1 ng in 2 µL when added to the master mix. The DNA stocks were initially subjected to a broad 1:10 dilution. Two representative strains (Dm28c_DTU-I and Perú_DTU-II) were then further diluted in a series of 1:2 dilutions to refine the limit of detection. The analytical limit of detection was defined as the lowest DNA concentration that resulted in positive amplification.

To calculate the equivalent number of genome/parasite copies, we used the formula:

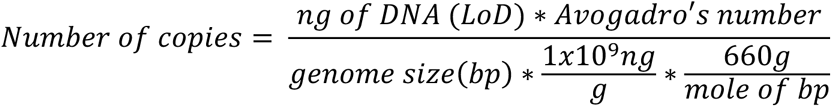

The genome size for nine strains (G, Dm28c, Tc1161, Y, Esmeraldo, SO3-cl5, CL, CL14, and CL Brener) was obtained from Souza et al., 2011^49^. The LoD was fixed in 0.00125 ng. The estimated number of copies was then divided by the median gene dose for *HSP70* in *T. cruzi*, which is 88, as determined using the gene copy number tool of TriTrypDB (https://tritrypdb.org/tritrypdb/app/search/transcript/GenesByCopyNumber).

### LAMP*_TcHSP70_* temperature stability

Twenty-four LAMP reactions were prepared and stored at −20°C. Four reactions were randomly selected, thawed, and subjected to amplification at six time points (months 0, 1, 2, 4, 6, and 8 post-storage). At each time point, three vials were loaded with 2 µL containing 0.5 ng of *T. cruzi* DNA (Dm28c_DTU-I), and the remaining vial was loaded with 2 µL of DNAse/RNAse-free water. The long-term stability was evaluated as changes in the mean fluorescence intensity using the ROI manager function of fiji2 software.

### One-Pot visual detection using Alt-R^TM^ CRISPR-Cas12a

The Cas12a-LAMP one-pot detection system was adapted from Qin et al., 2022. In a vial containing the previously described LAMP master mix, 2 µL of DNA template were added, followed by 35 µL of PCR-grade mineral oil. On top of the vial, 20 µL of the Cas12a reaction mix were added, which included: 2X NEB Buffer r2.1 (NEB, CN: B6002S), 8 U RNAase Inhibitor (ThermoFisher, CN: N8080119), 0.125 µM gRNA (5’-/AlTR1/rUrArArUrUrUrCrUrArCrUrArArGrUrGrUrArGrArUrCrGrUrCrArArUrGrCrGrCrUr CrGrCrGrCrUrGrU/AlTR2/-3’), 0.125 µM Alt-R™ L.b. Cas12a (Cpf1) Ultra (IDT, CN: 430891851), and 0.4 µM ssDNA probe (/56-FAM/TTATTATT/3IABkFQ/). The reaction was incubated at 65°C for 40 minutes. After incubation, the reaction was briefly spun down for 10 seconds to mix the Cas12a reaction mix with the LAMP amplified products. The mixture was then incubated again at 37°C for 20 minutes. Products were imaged under LED blue light using an Azure Biosystems 200 imaging system.

### Diagnostic test performance metrics

The sensitivity, specificity, and accuracy were estimated using the epiR (version 2.0.76) package in R, using a confidence level of 95%.

### Cost estimation for LAMP*_TcHSP70_* and TaqMan SatDNA qPCR

To calculate the total cost per reaction, the following categories were considered: reagents, disposable materials, personnel time, and permanent materials. All prices were based on costs in the United States. Annual depreciation for permanent materials was estimated using the formula: 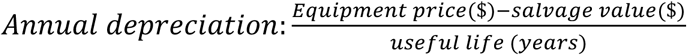

### *HSP70* amplicon sequencing

Parasites were isolated from patients in Santa Cruz Bolivia. The parasites were grown in culture for 3-4 months before cryopresevation. Parasites were then extracted with QIAmp DNA Blood Mini kit (QIAGEN, CN: 51104), and illumina sequencing libraries were prepared with NEBNext® Ultra™ II DNA Library Prep Kit (Illumina, E7645L) and barcoded with unique dual indexes for multiplexing. Libraries were sequenced in an Illumina NovaseqX with PE 150 bp reads. Reads aligned to *T. cruzi* were assembled using Spades v4.0.0^50^, with kmer lengths of 21,33,55,77,99, and 127 in paired-end mode. HSP sequences from these genomes were extracted from assemblies using a blast search against HSP70.

## ACKNOWLEDGMENTS

This work was supported by the National Institutes of Health R01AI151295 awarded to Dr. NB. We thank Dr. Daniel Urrea from the Universidad del Tolima in Colombia, and Dr. Juan David Ramirez from the Universidad del Rosario in Colombia for donating some of the DNA samples evaluated in this study.

## Data availability

All data produced in the present work are contained in the manuscript. A detailed protocol is available on protocols.io: DOI: dx.doi.org/10.17504/protocols.io.e6nvwbpqzvmk/v1. R code used to estimate the specificity and sensitivity is available on http://rpubs.com/sgutierr/1262176.

## Corresponding authors

Correspondence to Monica Mugnier

## Supplementary files

**Supplementary File 1:** SupFile1. Neighbor-Joining tree analysis and T. cruzi HSP70 identity.xlsx

**Supplementary File 2:** SupFile2. Primers sequences for the LAMP TcHSP70 assay.xlsx

**Supplementary File 3:** SupFile3. Representative alignment for HSP70 sequences.txt

**Supplementary File 4:** SupFile4. Heterodimer evaluation for LAMP TcHSP70 primers by Oligoanalyzer.xlsx

**Supplementary File 5:** SupFile5. Analytical limit of detection.xlsx

**Supplementary File 6:** SupFile6. Number of parasites equivalent to the LoD.xlsx

**Supplementary File 7:** SupFile7. Comparison of the limit of detection between the LAMP TcHSP70 assay and SatDNA qPCR.xlsx

**Supplementary File 8:** SupFile8. COST-estimation for LAMP and Taqman SatDNA qPCR.xlsx

**Supplementary File 9:** SupFile9. Mean fluorescence intensity after extended incubation and storing of LAMP TcHSP70 assay.xlsx

**Supplementary File 10.** SupFile10. qPCR and LAMP TcHSP70 results for clinical samples.xlsx

